# COVID-19 antibody seroprevalence in Duhok, Kurdistan Region, Iraq: A population-based study

**DOI:** 10.1101/2021.03.23.21254169

**Authors:** Nawfal R Hussein, Amer A Balatay, Ibrahim A Naqid, Shakir A Jamal, Narin A Rasheed, Alind N Ahmed, Reving S Salih, Ahmed S Mahdi, Sabeeha A Mansour, Shaveen Mahdi, Nashwan Ibrahim, Dildar H Musa, Zana SM. Saleem

## Abstract

**Objective:** This population-based study aimed to evaluate the seroprevalence of antibodies to SARS-CoV-2 in Duhok City, Kurdistan Region of Iraq.

**Methods:** We analyzed the national COVID-19 database that contains data regarding COVID-19 testing, management, and clinical outcomes in Duhok. For this study, different subdistricts within each district of Duhok were considered distinct clusters. Blood samples were collected from and questionnaires were administered to eligible and consenting participants who were members of different families from the subdistricts. Immunoassays were conducted to detect antibodies against SARS-CoV-2, and the associations between certain variables were investigated.

**Results:** The average number cases of COVID-19 before November 2020 was 23141 ± 4364, which was significantly higher than the average number of cases between November 2020 and February 2021 (3737 ± 2634; P = 0.001). A total of 743 individuals agreed to participate and were enrolled in the study. Among the participants, 465/743 (62.58%) were found to have antibodies against severe acute respiratory syndrome coronavirus 2. Among the participants with antibodies, 262/465 (56.34%) denied having any history of COVID-19-related symptoms. The most common symptom was fever (81.77%), followed by myalgia (81.28%). We found that antibody levels increased steadily with age (Pearson correlation coefficient = 0.117; P = 0.012). A significant association was found between antibody levels and the presence of symptoms (P = 0.023; odds ratio = 1.0023; 95% confidence interval = 1.0002-1.0061).

**Conclusions:** A significant reduction in the number of COVID-19 cases was observed. This might be due to the high prevalence of SARS-CoV-2 antibodies in Duhok. However, infection-prevention measures should be followed as it remains unclear whether acquired immunity is protective against reinfection. It expected that the infection rates during the next wave will not be as high as the first wave due to the high infection rate in the society.

## Introduction

Since the discovery of severe acute respiratory syndrome coronavirus 2 (SARS-CoV-2), the coronavirus disease (COVID-19) pandemic has been considered the most important healthcare crisis in the world. COVID-19 is an emerging infectious respiratory disease that was discovered in Wuhan, China, in December 2019 and rapidly spread worldwide [1]. The World Health Organization declared the COVID-19 outbreak a global pandemic on March 11, 2020. Since the appearance of the first few cases in Iraq, strict measures were put in place in the Kurdistan Region to stop the spread of infection [2]. The implementation of these measures began on March 1, 2020, and continued until May 21, 2020. After the measured were relaxed, a sharp increase in morbidity and mortality was recorded [3,4]. Despite this, there was an unwillingness in society to follow the social-distancing instructions [5]. In addition to a widely held belief that the COVID-19 pandemic is a hoax, education losses; widespread job losses; and food insecurity caused people not to adhere to the infection-containment measures [5]. The increase in morbidity and mortality started in mid-June 2020 and continued until October 2020. However, in November 2020, a significant reduction in the number of new COVID-19 cases was observed. This was unlikely to be the end of a wave of the COVID-19 pandemic in the Kurdistan Region because a majority of the residents in this area do not follow the prevention guidelines, and there are no specific measures being imposed by the Kurdistan Regional Government. One theory explaining this reduction in the number of cases is the achievement of herd immunity, which may play a role in slowing down the transmission of the virus. Herd immunity can be defined as the reduction in the number of infections or disease among the unimmunized individuals of a population as a result of previous infection or immunization in a proportion of individuals of the population [6]. It can be determined by testing a sample of a population for the presence of immune markers [6]. The aim of this population-based study was to evaluate the seroprevalence of antibodies to SARS-CoV-2 among people in Duhok City.

## Materials and methods

### Data source

For a national project, all data regarding COVID-19 testing, management, and clinical outcomes in Duhok were reported and registered in a COVID-19 database. The COVID-19 database contains information regarding all the confirmed cases of COVID-19 in the city, showing the results of reverse-transcription polymerase chain reaction testing. In addition, the database covers the number of tests performed, number of suspected cases, cure rate, case fatality rates, and clinical outcomes. The data were analyzed to confirm the decline in the number of cases in the city.

### Study sampling

Regional representation was achieved by dividing the city into five districts, and sampling was performed based on population proportions. Because of considerable differences in population density among the different geographic regions of Duhok, we included samples from both urban and rural populations. Random samples were selected using multistage cluster sampling. First, within each district, different subdistricts were considered to be distinct clusters. Subdistricts are known regions within districts with clear boundaries of the city. Second, housing censuses of households in the selected clusters were conducted by a sample-collecting team. General information on the number of families and family members was collected. Then, a number of families within each region were chosen and asked whether they would agree to participate. If a family did not agree to participate, they were excluded from the study. Finally, from each selected family, two people were randomly chosen to participate. The people who did not agree to participate or those who were absent on the day of sample collection were excluded from the study and replaced with other participants.

### Sample size

When we started the study, an increasing number of unpublished studies reported that the prevalence of COVID-19 had reached 50% in Duhok City, and because there had been no previous reports of the prevalence of the infection in the city, a prevalence of 50% was used to calculate the sample size. With a confidence interval of 99%, margin of error of 0.05, design effect of 1, and expected response rate of 80%, the minimum sample size calculated was 830.

### Inclusion criteria

A person was eligible to be included in the study if the person was at least 16 years old, was a resident of one of the considered districts, had been living at a specific address for at least 6 months, and agreed to participate in the study.

### Sample collection

Samples were collected between January 10, 2021, and January 30, 2021. A week before the survey, the team members who had not been trained on the use of personal protective equipment attended a training session on their use. All the team members of the sample-collection team attended a training session regarding the blood-sampling process employed in this study and the administration of a questionnaire. The persons who were eligible for inclusion in the study were interviewed and they completed a questionnaire. Subsequently, a member of the team explained the procedures for sample collection. Then, 5-10 cm^3^ of venous blood samples were collected using 10-cm^3^ syringes. The samples were immediately transported to the research center, and sera were separated from the blood and kept frozen at −20°C.

### SARS-CoV-2 antibody measurement

Elecsys Anti-SARS-CoV-2 (Roche Diagnostics International Ltd, Rotkreuz, Switzerland), which is an immunoassay for the in vitro qualitative detection of antibodies (including IgG) to SARS-CoV-2, was used to determine antibody levels. The assay was performed according to the manufacturer’s instructions. According to the manufacturer, a cutoff index ≥ 1.0 indicates a positive result.

### Statistics

When variables were continuous, linear regression was used to study the relationship between the variable and outcome. A Chi-square test was used to analyze categorical data. Pearson’s correlation coefficients were calculated to study the relationship between two continuous variables. Statistical analysis was performed using Minitab 17. P values < 0.05 were considered significant.

### Ethics

The study protocol and method of obtaining consent were approved by the Ethics and Scientific Committee of the University of Zakho’s College of Medicine. Informed consent was obtained from all participants.

## Results

### Data analysis

The analysis of the national COVID-19 database showed a sharp decrease in the percentage of new cases (Fig 1). The average number of COVID-19 cases before November 2020 was 23141 ± 4364, which was significantly higher than the average number of cases between November 2020 and February 2021 (3737 ± 2634; P = 0.001).

**Fig 1.**
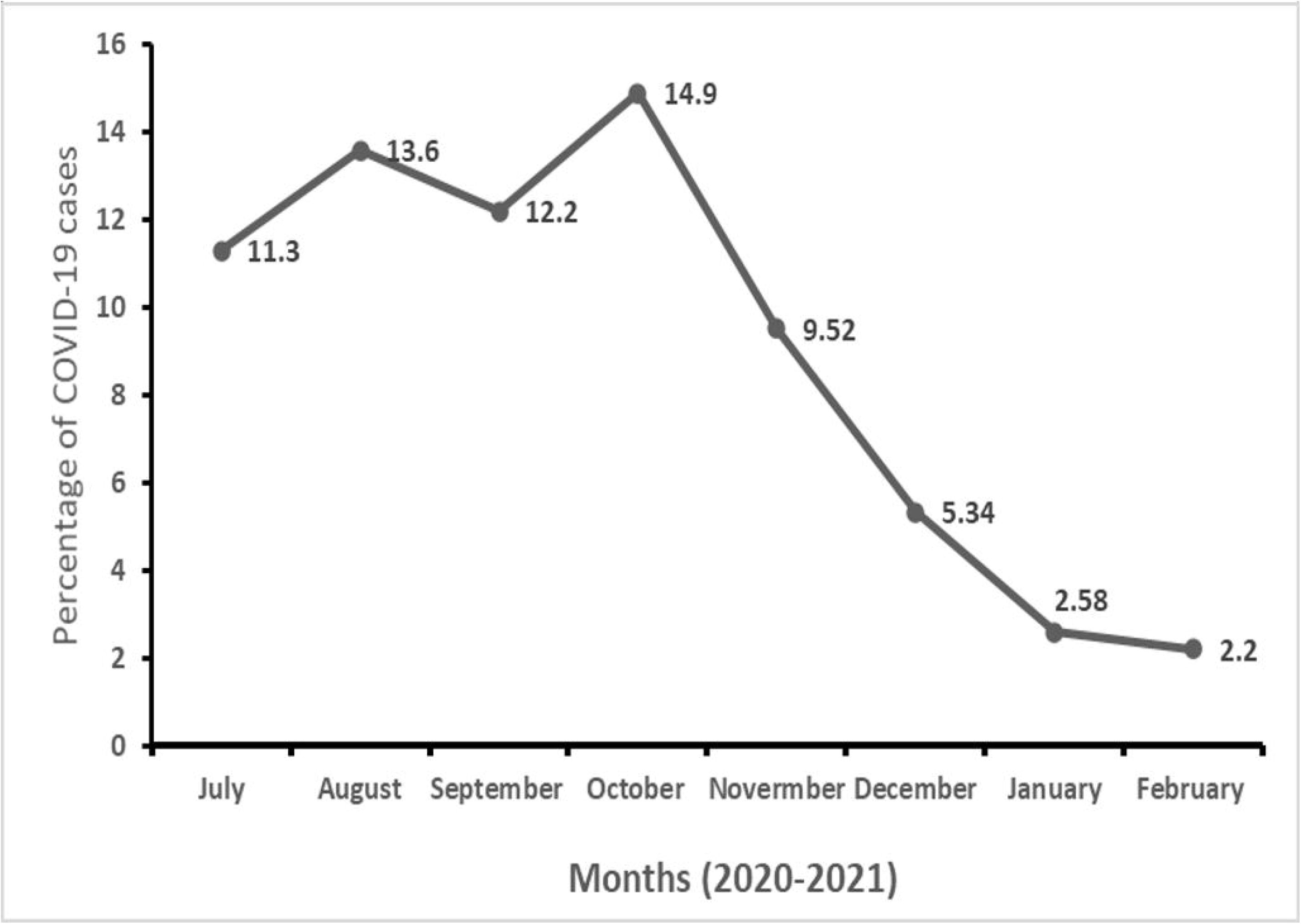
Reduced number of COVID-19 cases. Significant reduction in the number of cases in Duhok, based on the incidence of reported cases in the city.

### Prevalence of COVID-19

A total of 903 individuals were interviewed in this study, which considered 411 households in five districts of Duhok; 192 individuals lived in the city center, and the rest were from peripheral towns and villages. In this study, 160 individuals did not agree to participate and were excluded from further analysis. A total of 743 individuals agreed to participate and were enrolled in the study. The average age of the participants was 34.6 ± 13.9, and (392/743) 52.8% of the participants were female. Among the participants, 519/743 were married and (164/743) 22.1% had a history of chronic diseases.

### Antibody positivity and association between host factors and antibody levels

Among the participants, 465/743 (62.58%) tested positive for antibodies. Among the participants with antibodies, 262/465 (56.34%) denied having any history of COVID-19 related symptoms. A total of 203/465 (43.66%) participants had a history of different symptoms (Table 1). The average duration of symptoms was 11.81 ± 10.80 days, and the most common symptom was fever (81.77%), followed by myalgia (81.28%) and loss of smell (56.79%). Among the participants who had experienced symptoms, 21.67% reported having experienced diarrhea (Table 1). The presence of symptoms was significantly higher among married participants (P=0.046), participants with a history of chronic diseases (P = 0.001), and older participants (P = 0.0015) (Table 2).

**Table 1.** Characteristics of participants who tested positive for antibodies.

| Characteristics |  | No. of participants | % |
| --- | --- | --- | --- |
| Sex: Female |  | 264 | 56.77 |
| Marital status: Married |  | 335 | 72.04 |
| Chronic diseases (Yes) |  | 113 | 24.3 |
| Symptoms |  | 203 | 43.87 |
| Symptoms | Fever | 166 | 81.77 |
|  | Myalgia | 165 | 81.28 |
|  | Loss of smell | 114 | 56.79 |
|  | Loss of taste | 95 | 46.79 |
|  | Shortness of breath | 89 | 43.84 |
|  | Cough | 41 | 20.19 |
|  | Diarrhea | 44 | 21.67 |
|  | Headache | 40 | 19.7 |
| Duration of symptoms (Days + standard deviation) |  | 11.81 ± 10.80 |  |
| Age |  | 34.40 ± 13.73 |  |

**Table 2.**
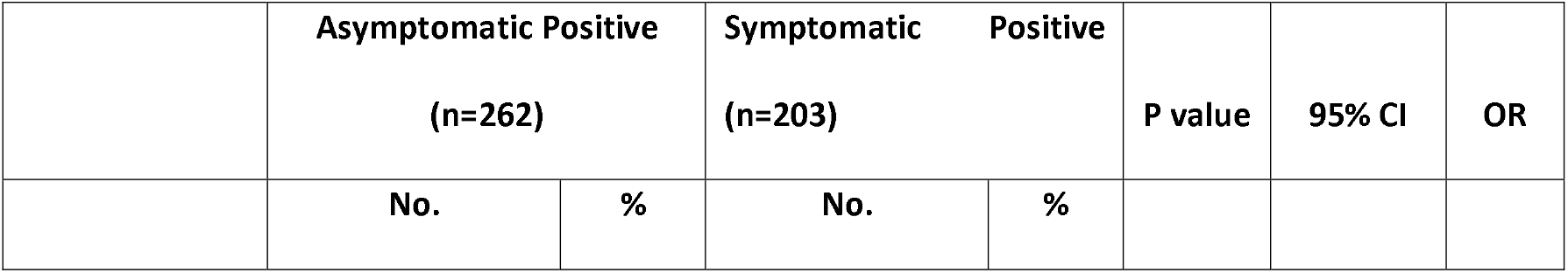
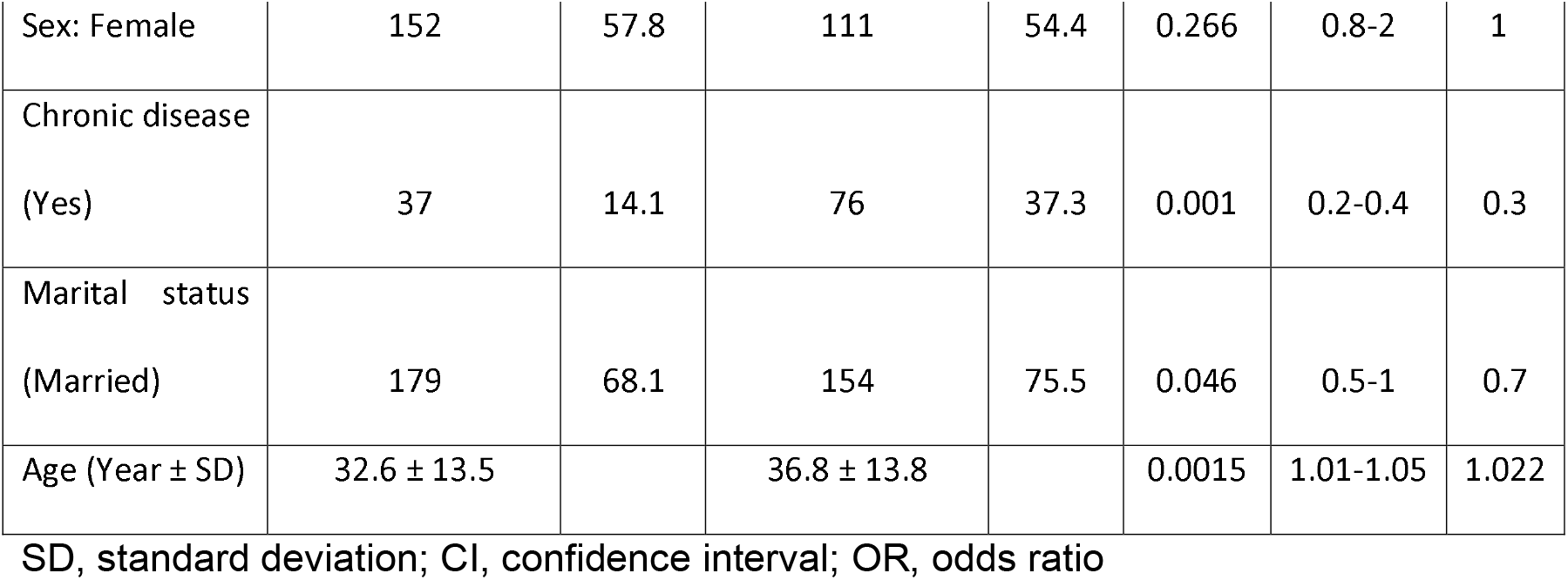
Associations between different factors and antibody levels.

|  | Asymptomatic Positive<br>(n=262) |  | Symptomatic Positive<br>(n=203) |  | P value | 95% CI | OR |
| --- | --- | --- | --- | --- | --- | --- | --- |
|  | No. | % | No. | % |  |  |  |

**Table 2. Associations between different factors and antibody levels**
|  |  |  |  |  |  |  |  |
| --- | --- | --- | --- | --- | --- | --- | --- |
| Sex: Female | 152 | 57.8 | 111 | 54.4 | 0.266 | 0.8-2 | 1 |
| Chronic disease<br>(Yes) | 37 | 14.1 | 76 | 37.3 | 0.001 | 0.2-0.4 | 0.3 |
| Marital status<br>(Married) | 179 | 68.1 | 154 | 75.5 | 0.046 | 0.5-1 | 0.7 |
| Age (Year ± SD) | 32.6 ± 13.5 | | 36.8 ± 13.8 | | 0.0015 | 1.01-1.05 | 1.022 |

SD, standard deviation; CI, confidence interval; OR, odds ratio

Pearson’s correlation coefficient was calculated to explore the relationship between antibody levels and the age of the participants. We found that antibody levels increased steadily with age (Pearson’s correlation coefficient = 0.117; P = 0.012) (Fig 2A). No association was found between the time interval between SARS-CoV-2 detected and testing date (Pearson’s correlation coefficient = 0.029; P = 0.681) (Fig 2B). Linear regression analysis was used to investigate antibody levels, sex, marital status, and the presence of symptoms (Table 2). A significant association was found between antibody levels and the presence of symptoms (P = 0.023; odds ratio = 1.0023; 95% confidence interval = 1.0002-1.0061). No significant associations were found between antibody levels and sex or marital status (Table 2).

**Fig 2.**
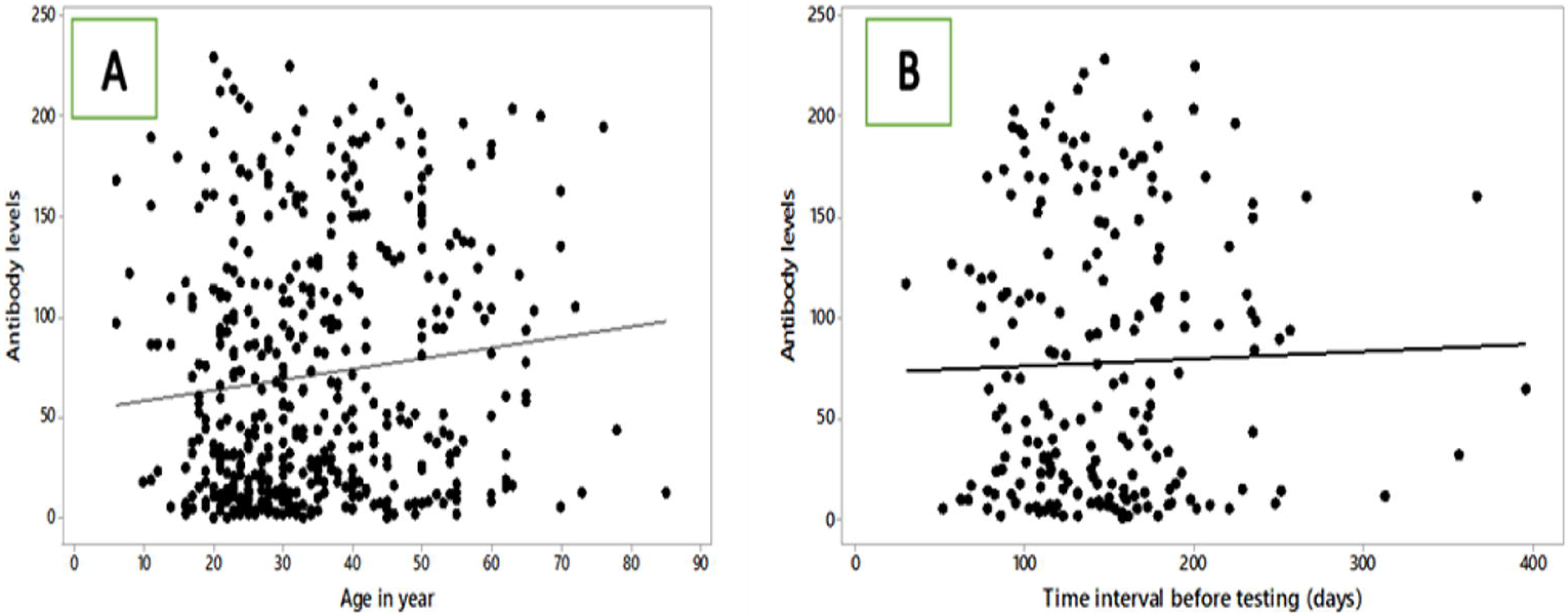
Association of age and time interval before testing with antibody responses to SARS–CoV-2. (A) Age was associated with antibody responses to SARS-CoV-2. Analysis through calculation of Pearson’s correlation coefficient was used to investigate the relationship between age and antibody levels. (B) Time interval before testing was not associated with antibody responses to SARS-CoV-2. Analysis through calculation of Pearson’s correlation coefficient was used to investigate the relationship between time intervals and antibody levels.

## Discussion

Since the emergence of the COVID-19 pandemic, scientific communities worldwide have aimed to control the spread of the infection. The achievement of herd immunity has been highlighted as an option for controlling the pandemic. Certain questions remain regarding whether herd immunity is derived from natural infection and what proportion of a population must attain immunity through infection for the effect of protection to be achieved. Since the basic reproduction number for COVID-19 has been estimated to lie within the range of 2-4, the herd immunity level for COVID-19 is believed to lie between 50% and 75% [7]. In one study, the authors assumed that that all individuals are equally susceptible and equally infectious, and the threshold to achieve herd immunity of SARS-CoV-2 was calculated to range between 50% and 67% [8]. In a study conducted in Bangladesh, the computation model indicated that the herd immunity threshold can be reduced from 60% to 31% by categorizing the population into age groups [9]. In Duhok, strict measures were imposed by the government to control the spread of the infection [2]; however, mounting economic and political pressures pressurized the government into relaxing the measures. The relaxation of the measures took place at a rapid pace, without any specific plans [5]. The government’s actions in this regard were then limited to the giving of instructions, and those who did not follow the instructions were not penalized. The number of COVID-19 cases increased sharply until November 2020, when a sharp decrease in the number of cases was observed [3,4]. Our study shows that the number of cases decreased significantly in the following months. This is unlikely to be the end of a wave because no specific measures were taken, and people did not follow the social-distancing instructions [3,5]. We aimed to investigate this decline in the number of cases. In this study, the prevalence of COVID-19 was found to be > 62%. Most significantly, in the present study, the number of infections was found to be much greater than the registered number of cases by the Goverment, which was approximately 3% [10]. This finding implies that there were approximately 20 cases in Duhok City for each registered case. Thus, the number of registered cases was 20-fold lower than the number of COVID-19 cases predicted by this study. This may be due to a great number of individuals with infection being asymptomatic or cases not being reported because of the stigma associated with the infection. This ascertainment and difference between our results and registered cases is a crucial because infection-to-case ratios are considered in many projection and epidemiological models in order to calculate fatality rates and understand which stage an epidemic is in [11]. Additionally, such an infection-to-case ratio may suggest that the fatality rate might be much higher. Another important implication of this study’s findings is that with such a high prevalence, it is unlikely that the number of cases will increase sharply again to previous levels recorded during the first wave. Since antibody titers may decline after a period of time, the prevalence of SARS-CoV-2 infection may be higher than what we reported. It is worth mentioning that it is unclear whether post-infection immunity, specifically that provided by antibodies, protects individuals or generates herd immunity, and further studies are required to explore this. Furthermore, more than 56% of the participants who tested positive for antibodies were asymptomatic. Although the exact role of asymptomatic individuals in the transmission of the virus is not clear [12], the great number of asymptomatic individuals with SARS-CoV-2 infection may have played a role in the widespread transmission of the virus in Duhok City.

Next, we studied the factors that may impact the production of antibodies. In agreement with previous reports [13,14], we found that older age was associated with higher antibody levels. This may be because older individuals are more susceptible to severe infection. In contrast to a previous report [15], no association was found between sex and antibody levels. In agreement with a previous study [13,16], we found an association between the presence of symptoms and higher antibody levels. Further investigation is needed to study the effects of different factors and their association with antibody levels. It has been previously shown that antibody levels may decline over time [17,18]. In our study, no association was found between the time interval before testing and antibody levels. This may be partially explained by differences in human genetics, virus genetic makeup, and environmental factors. It is worth mentioning that our study was cross-sectional; therefore, the kinetics of antibody response over time could not be investigated.

Our study had certain limitations. First, we could not ascertain the representativeness of SARS-CoV-2 antibodies in groups with possibly high prevalence, such as healthcare workers and nursing-home residents and workers. Additionally, we could not reach remote villages due to transportation difficulties, particularly those caused by the snowy winter weather.

In conclusion, a significant reduction in the number of COVID-19 cases was observed. This may be due to the high prevalence of SARS-CoV-2 antibodies in Duhok. To control the spread of COVID-19, it is extremely important for members of society to follow social-distancing measures. It expected that the infection rates during the second wave will not be as high as the first wave due to the high infection rate in the society. Locally derived population prevalence estimates should be used to develop infection control plans.

## Data Availability

All data will be available, if requested, through the email of corresponding author.

## Acknowledgments

We would like to thanks Amer Lab staff for the technical support. We Acknowledge the role of Mr Muhammed Salih for helping in recruiting participants into the project.

